# Longitudinal analysis of laboratory findings during the process of recovery for patients with COVID-19

**DOI:** 10.1101/2020.04.04.20053280

**Authors:** Suyan Tian, Xuetong Zhu, Xuejuan Sun, Jinmei Wang, Qi Zhou, Chi Wang, Li Chen, Jiancheng Xu

## Abstract

**Objective:** To explore longitudinal change patterns of key laboratory tests in patients with COVID-19, and to identify independent prognostic factors by examining the associations between laboratory findings and outcomes of patients.

**Methods:** The multicenter study prospectively included 59 patients with COVID-19 treated at Jilin province from January 21, 2020 to May 5, 2020. Laboratory tests were included haematological, biochemical, and immunological tests.

**Results:** Laboratory findings, the characteristics of epidemiological and demographic data were extracted from electronic medical records. Eosinopenia was shown in 52.6% cases at onset, and the average value of eosinophil continued to significantly increase thereafter. Lymphopenia was found in 40.4% cases at onset, and the average value of lymphocyte was slowly elevated after day 5. Thrombocytopenia was shown in 12.3% cases at onset, and the average value of mean platelet volume was decreased sharply after day 7. The values of aspartate aminotransferase, lactate dehydrogenase, creatine kinase, creatinine kinase-muscle/brain activity, and cardiac troponin I, serum cardiac markers, were beyond the upper limit of RI from 6.1% to 30.6% at onset. The abnormity of liver function tests, kidney function tests, electrolytes was 2.0%∼59.2%, 2.0%∼4.1%, 6.0%∼30.0%, respectively. Eosinophil, platelet and carbondioxide combining power were selected as the prognostic factors.

**Conclusions:** The haematological, biochemical, and immunological tests were found significant abnormity at onset and longitudinal change patterns in the patients with COVID-19. Age, Eosinophil, PLT and CO_2_ may used to predict the recovery probability. Risk stratification and management could be improved for the patients with COVID-19 according to temporal trajectories of laboratory tests.

**Article Summary:** The longitudinal change patterns of the laboratory characteristics of affected patients were important to identify prognosis. Eosinophil, platelet and carbondioxide combining power may be independent predictors of recovery in patients with COVID-19.

## Introduction

Since the emergence of Corona virus disease 2019 (COVID-19) caused by severe acute respiratory syndrome coronavirus 2 (SARS-CoV-2) in Wuhan, China, in December 2019,(1) it has rapidly spread across China and many other countries.(2) Mild type COVID-19 was found in most patients with no obvious abnormal CT findings or manifestation of lung infection. Patients with common type often presented fever, respiratory symptoms and abnormal CT findings. Severe type COVID-19 was designated when the patients had one of the following criteria: 1) Respiratory distress with respiratory frequency ≥ 30/min; 2) Oxygen Saturation ≤ 93% at rest; 3) Oxygenation index (artery partial pressure of oxygen/inspired oxygen fraction, PaO_2_/FiO_2_) ≤ 300 mmHg. However, patients with critical type progressed rapidly to acute respiratory failure, acute respiratory distress syndrome, metabolic acidosis,coagulopathy, and septic shock. In clinical practice, it is found that some patients’ condition will suddenly worsen, which brings challenges to the treatment. Early identification of risk factors for severe illness facilitated appropriate treatment and prognosis. Therefore, it is necessary to make a comprehensive judgment with laboratory findings through the patient’s condition.

Although a few studies have reported epidemiologic features, molecular characteristics, clinical manifestations, chest images, and laboratory findings of the patients with COVID-19,(3, 4)these studies are most of cross-sectional studies and cannot reflect the whole course of the disease. To the best of our knowledge, comprehensive investigation on dynamic change trajectories over time of multiple laboratory data in recovered patients with COVID-19 has not yet been reported. The temporal change patterns of laboratory data may provide insightful clues on the whole course of the disease. Therefore, it is essential to identify longitudinally differential laboratory characteristics in patients with COVID-19.

The present research aimed to investigate longitudinal change patterns of laboratory data in recovered patients with COVID-19 and reveal the associations between laboratory findings and outcomes of patients to provide prognostic factors from laboratory findings.

## Methods

### Patient selection and data sources

We performed an observational study on the laboratory characteristics of confirmed patients with COVID-19. The confirmed case with COVID-19 was defined as a positive result to real-time reverse-transcriptasepolymerase-chain-reaction (RT-PCR) assay for sputum and throat swabs pecimens.(1) In this multi-center study, 59 patients were recruited in the multicenter study from three designated tertiary hospitals in Jilin province including the First Hospital of Jilin University (*n*=3), Changchun Infectious Disease Hospital (*n*=42), and Siping Infectious Disease Hospital (*n*=14) from January 21, 2020 to March 5, 2020. Laboratory findings, the characteristics of epidemiological and demographic data were retrieved from electronic medical records. Two researchers independently checked the data. The incubation period was defined as the duration from the contact of the transmission source to the onset of symptoms. The date of last discharged patient was March 5, 2020, the final date of follow-up. The study was approved by the Ethics Committee of the First Hospital of Jilin University, Changchun Infectious Disease Hospital and Siping Infectious Disease Hospital. Written informed consent was waived in review of the emergency need to collect clinical data.

### Laboratory tests

Laboratory confirmation of COVID-19 was achieved through Changchun Center for Disease Control and Prevention, or Siping Center for Disease Control and Prevention. Suspicious positive sputum and throat swab specimens were transported to confirm again by Jilin Provincial Center for Disease Control and Prevention for laboratory diagnosis. Specimens were tested by RT-PCR for SARS-Cov-2 RNA. The RT-PCR assay was conducted in accordance with the manufacturer’s protocol (Shanghai bio-germ Medical Technology Co. Ltd, and Shanghai GeneoDx Biotech Co. Ltd). Patients with COVID-19 were recovered from hospital once the results of two RT-PCR tests taken 24 hours apart were negative for SARS-Cov-2.

Laboratory assessments including haematological, biochemical, and immunological tests were determined in the three designated tertiary hospitals. Test kits, calibrators, and quality control products were matched same equipment. All doctors, technicians, and nurses in this study received unified training by Health Commission of Jilin Province. The Ministry of Health of the People’s Republic of China published common biochemical analyte and blood cell analysis reference intervals of Chinese adults from 2012.(5, 6)These reference intervals of items in the three designated tertiary hospitals were all used by these China Industry Standards.

### Statistical analysis

In order to estimate temporal trajectories of primary markers, local regression curves (loess) were diagramed by using R locfit package. The corresponding 95 % confidence intervals of the estimated values at days 0-30 were given, with the aids of the bootstrapping methods (here, 1000 bootstrapped data were made).

To identify tests that have prognostic values to predict patient’s recovery time, univariate Cox models were firstly fit. Upon the markers that have *P*-values < 0.1 in the univariate analysis, gender and age, three feature selection methods — the LASSO method, multivariate Cox regression with backward selection and multivariate Cox regression with stepwise selection were used to further refine the list. For the final model, a nomogram was made to illustrate how the selected markers were associated with the chance of recovery. All analysis were carried out using the R language, version 3.6.1 (www.r-project.org).

## Results

### Demographics and exposures

Of all 59 cases diagnosed as COVID-19 in the three designated tertiary hospitals in Jilin province, 56 were diagnosed as common type and 3 as severe type. The median age was 41 years, ranging from 10 to 87 years old (IQR: 29-52), and 27.1% (16/59) were more than 50 years old. 42.4% (25/59) of cases were female. The median intervals of incubation period were 4 days (IQR, 2-7). At the final date of follow-up, all the 59 patients were discharged. The average length of hospital stay was 17 days (IQR, 11-20). 15.3% (9/59) of cases had travel history to Wuhan, and 28.8% (17/59) had travel history outside Wuhan. 42.3% (25/59) were primary cases, who had not left Jilin province recently, but had a close exposure history with imported cases, and 13.6% (8/59) were secondary cases, who had a close exposure history with primary contacts. The transmission route for COVID-19 is shown in Figure 1. The baseline characteristics of recovered cases with COVID-19 were shown in Table 1.

**Table 1.** The baseline characteristics of recovered cases with COVID-19.

|  | Male (*n*=34) | Female (*n*=25) |
| --- | --- | --- |
| **Hospitalized time, d, M (P<sub>25</sub>-P<sub>75</sub>)** | 17 (13-21) | 17 (11-19) |
| **Age, y, M (P<sub>25</sub>-P<sub>75</sub>)** | 41 (29-51) | 41 (28-53) |
| **Travel history in Wuhan** | 7 (20.6) | 2 (8.0) |
| **City** |  |  |
| Changchun | 25 (73.5) | 20 (80.0) |
| Siping | 9 (26.5) | 5 (20.0) |
| **Age groups** |  |  |
| <30y | 10 (29.4) | 8 (32.0) |
| 30-49y | 15 (44.1) | 10 (40.0) |
| 50-69y | 6 (17.7) | 5 (20.0) |
| ≥70y | 3 (8.8) | 2 (8.0) |
| **Contact category** |  |  |
| Primary cases | 17 (60.7) | 9 (36.0) |
| Secondary cases | 11 (28.6) | 14 (56.0) |
| Tertiary cases | 6 (10.7) | 2 (8.0) |

Data are n (%) unless specified otherwise. y: year; d: day.

**Figure 1.**
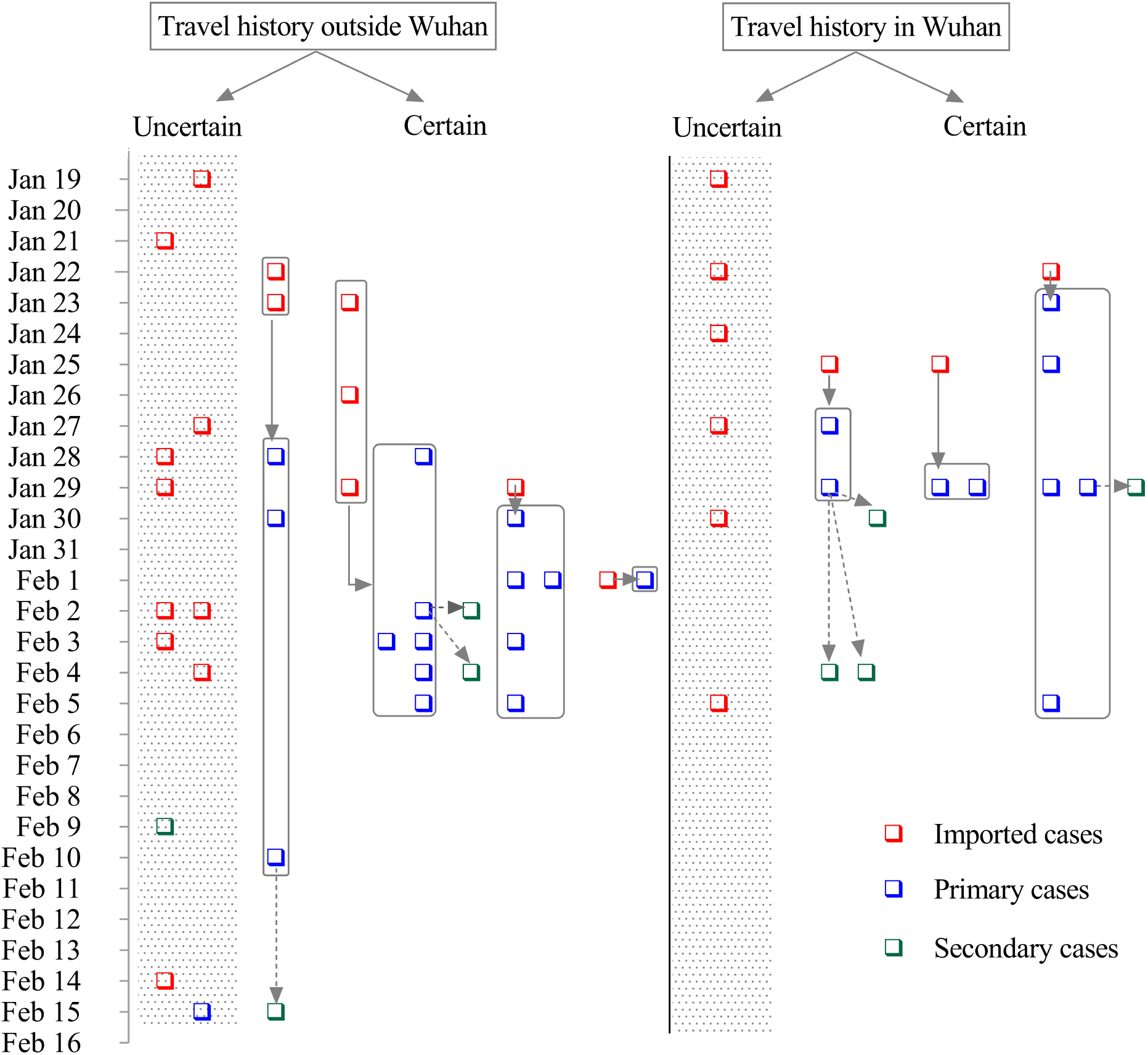
Transmission route for patients with COVID-19 in Jilin province. The route is plotted according to the time of onset. Travel history: travel history of imported cases; Squares: confirmed cases; Imported cases: patients who had travel history in Hubei Province or contact history with confirmed patients outside Jilin Province within 14 days before the onset of symptoms; Primary cases: patients caused by imported cases; Secondary cases: patients caused by primary cases; Uncertain: patients who are not link to any specific individual.

### Laboratory findings at onset to hospital admission

As leukocyte parameters, the counts of white blood cell (WBC), neutrophils (NE), lymphocytes (LY), and eosinophils (EO) below the lower limit of RI accounted for 14.0%∼52.6%, while both basophils (BA) and monocytes (MO) beyond the upper limit of RI accounted for 5.3% at onset. As erythrocyte parameters, decreased counts of red blood cells (RBC), hemoglobin (HGB), hematocrit (HCT), mean corpuscular volume (MCV) and mean corpuscular hemoglobin (MCH) accounted for 1.8%∼12.3%. As platelet parameters, the counts of platelets count (PLT) and the value of thrombocytocrit (PCT) below the lower limit of RI accounted for 12.3% and 5.3%, respectively. As coagulation parameters, the values of activated partial thromboplastin time (APTT), prothrombin time (PT), and international normalized ratio (INR) above the upper limit of RI were found in 8.1%∼24.3% cases, while the values of fibrinogen (FBG) below the lower limit of RI were found in 16.2% cases. The concentration of high-sensitivity C-reactive protein (hsCRP) was higher than the upper limit of RI in 51.1% cases. As serum cardiac markers, the values of aspartate aminotransferase (AST), lactate dehydrogenase (LDH), creatine kinase (CK), creatinine kinase-muscle/brain activity (CK-MB), and cardiac troponin I (cTnI) beyond the upper limit of RI accounted for 6.1%∼30.6%. As liver function tests, the values of alanine aminotransferase (ALT), γ-glutamyl transpeptidase (GGT), total bilirubin (TBIL), and direct bilirubin (DBIL) above the upper limit of RI accounted for 2.0%∼26.5%, while the values of cholinesterase (CHE), total protein (TP), albumin (ALB), globulin (GLB), and indirect bilirubin (IBIL) below the lower limit of RI accounted for 2.4%∼59.2%. As kidney function tests, the values of blood urea nitrogen (BUN) and creatinine (Cr) above the upper limit of RI accounted for 2.0%, and 4.1%, respectively. As electrolyte tests, potassium (K), sodium (Na) and chloride (Cl) below the lower limit of RI accounted for 6.0%∼30.0%, respectively. There were gender differences in RBC, HGB, HCT, MCV, MCH, mean corpuscular hemoglobin concentration (MCHC), PCT, thrombin time (TT), PT, CK, GGT, TBIL, DBIL, Cr, and Cl. Laboratory findings of patients with COVID-19 at onset to hospital admission were shown in Table 2.

**Table 2.**
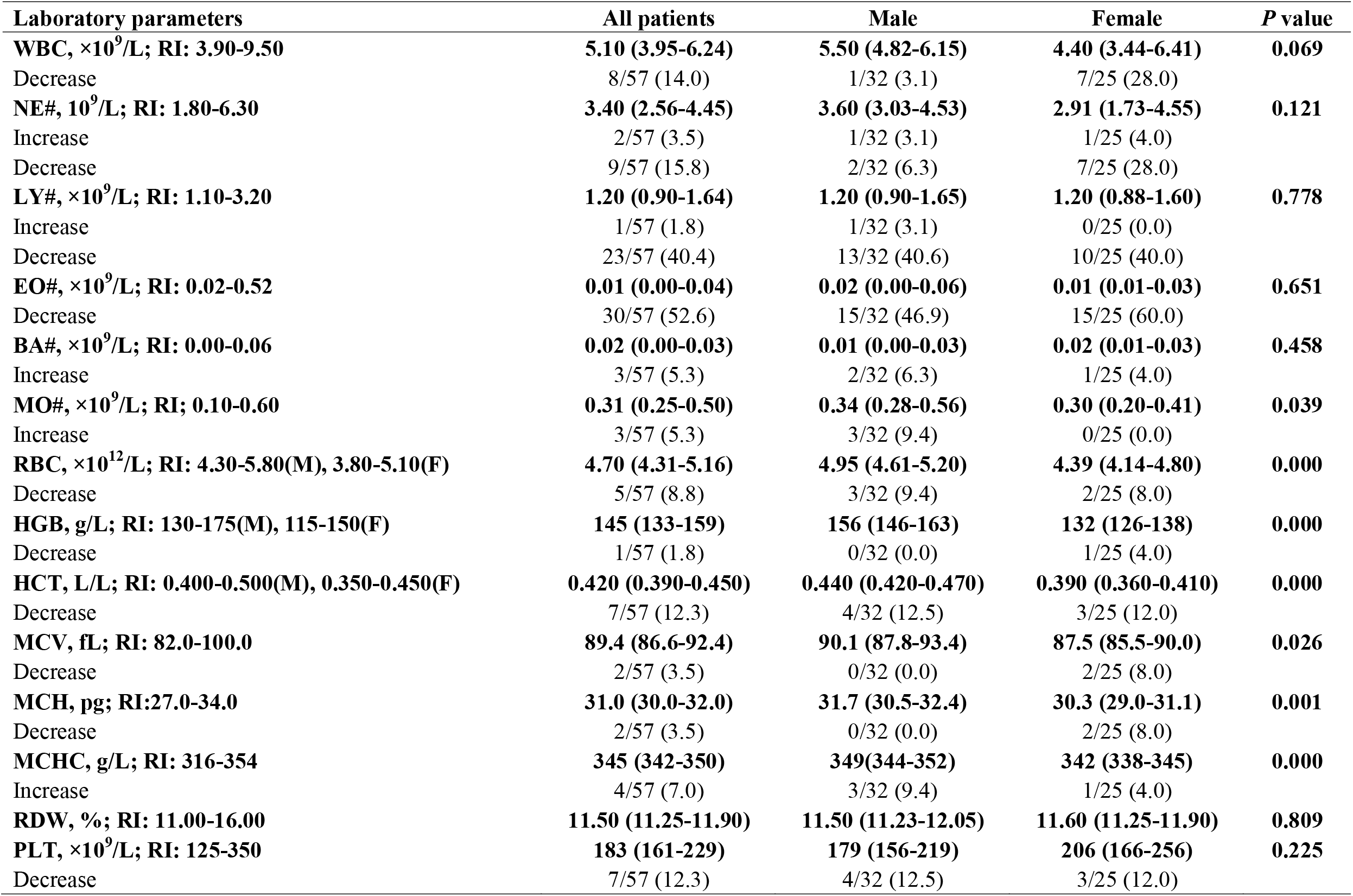
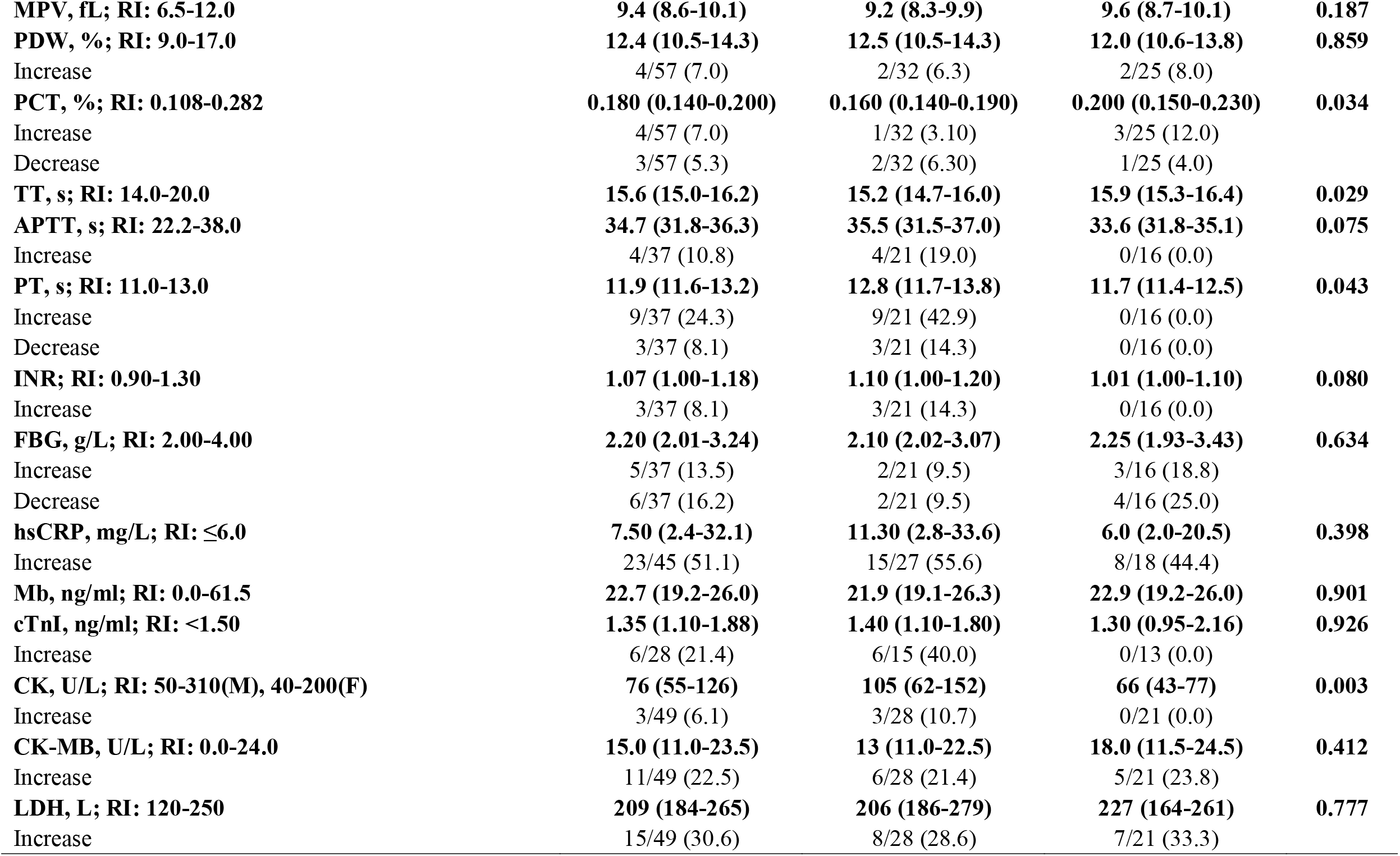
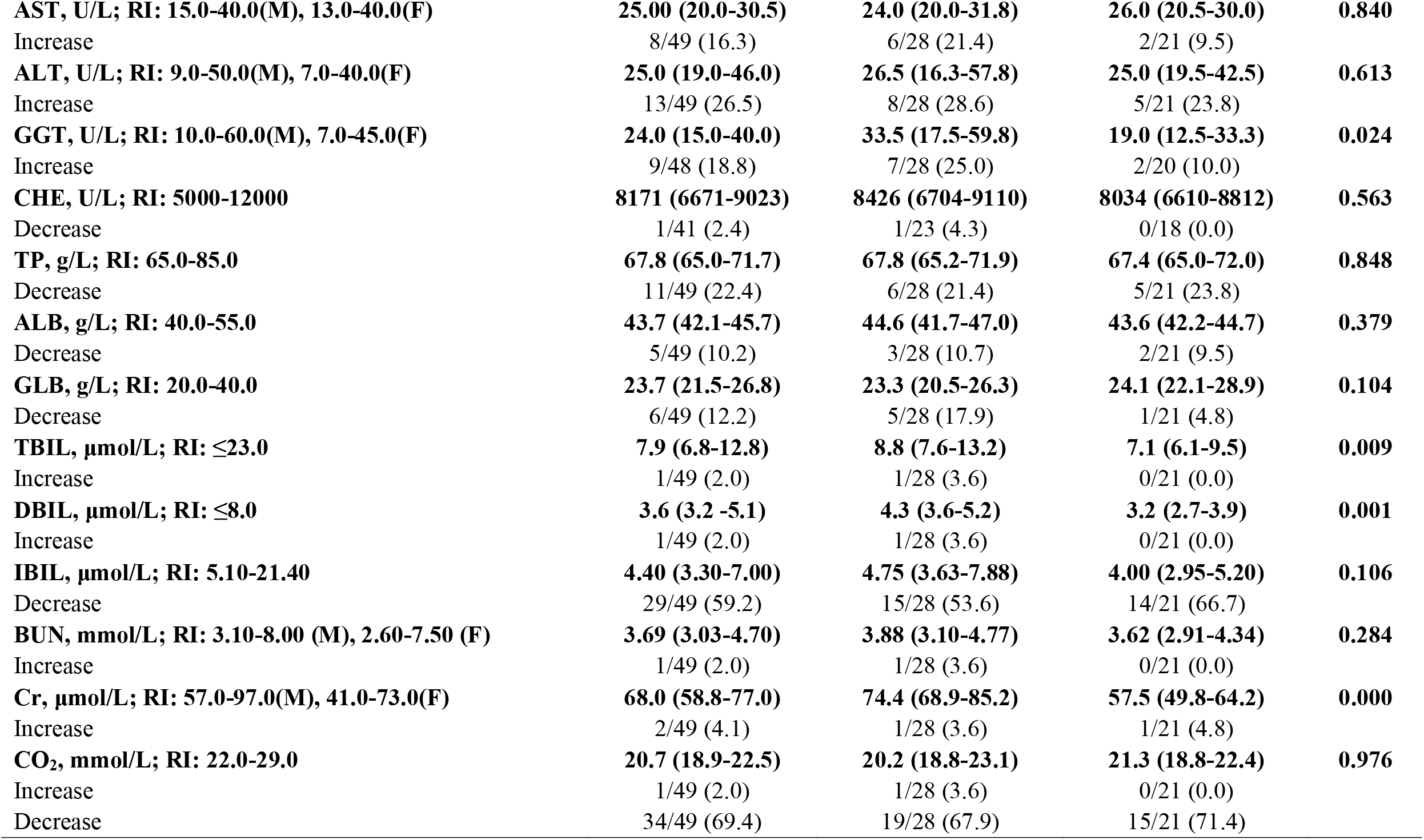
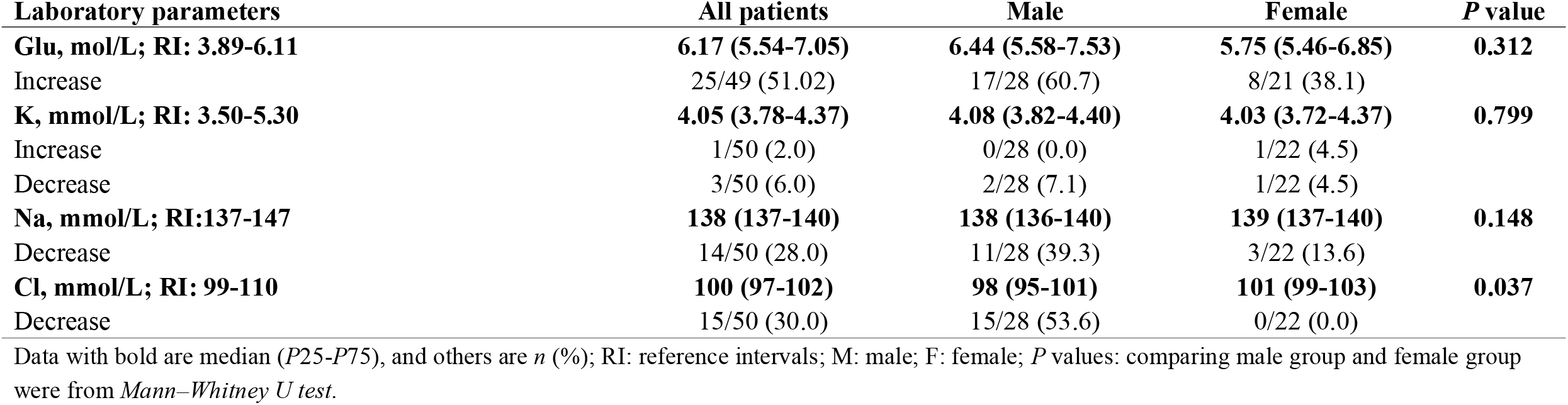
Laboratory findings of patients with COVID-19 at onset to hospital admission.

| **Laboratory parameters** | **All patients** | **Male** | **Female** | **P value** |
| --- | --- | --- | --- | --- |
| **WBC, $\times 10^9/L$; RI: 3.90-9.50** | **5.10 (3.95-6.24)** | **5.50 (4.82-6.15)** | **4.40 (3.44-6.41)** | **0.069** |
| Decrease | 8/57 (14.0) | 1/32 (3.1) | 7/25 (28.0) |  |
| **NE#, $10^9/L$; RI: 1.80-6.30** | **3.40 (2.56-4.45)** | **3.60 (3.03-4.53)** | **2.91 (1.73-4.55)** | **0.121** |
| Increase | 2/57 (3.5) | 1/32 (3.1) | 1/25 (4.0) |  |
| Decrease | 9/57 (15.8) | 2/32 (6.3) | 7/25 (28.0) |  |
| **LY#, $\times 10^9/L$; RI: 1.10-3.20** | **1.20 (0.90-1.64)** | **1.20 (0.90-1.65)** | **1.20 (0.88-1.60)** | **0.778** |
| Increase | 1/57 (1.8) | 1/32 (3.1) | 0/25 (0.0) |  |
| Decrease | 23/57 (40.4) | 13/32 (40.6) | 10/25 (40.0) |  |
| **EO#, $\times 10^9/L$; RI: 0.02-0.52** | **0.01 (0.00-0.04)** | **0.02 (0.00-0.06)** | **0.01 (0.01-0.03)** | **0.651** |
| Decrease | 30/57 (52.6) | 15/32 (46.9) | 15/25 (60.0) |  |
| **BA#, $\times 10^9/L$; RI: 0.00-0.06** | **0.02 (0.00-0.03)** | **0.01 (0.00-0.03)** | **0.02 (0.01-0.03)** | **0.458** |
| Increase | 3/57 (5.3) | 2/32 (6.3) | 1/25 (4.0) |  |
| **MO#, $\times 10^9/L$; RI: 0.10-0.60** | **0.31 (0.25-0.50)** | **0.34 (0.28-0.56)** | **0.30 (0.20-0.41)** | **0.039** |
| Increase | 3/57 (5.3) | 3/32 (9.4) | 0/25 (0.0) |  |
| **RBC, $\times 10^{12}/L$; RI: 4.30-5.80(M), 3.80-5.10(F)** | **4.70 (4.31-5.16)** | **4.95 (4.61-5.20)** | **4.39 (4.14-4.80)** | **0.000** |
| Decrease | 5/57 (8.8) | 3/32 (9.4) | 2/25 (8.0) |  |
| **HGB, g/L; RI: 130-175(M), 115-150(F)** | **145 (133-159)** | **156 (146-163)** | **132 (126-138)** | **0.000** |
| Decrease | 1/57 (1.8) | 0/32 (0.0) | 1/25 (4.0) |  |
| **HCT, L/L; RI: 0.400-0.500(M), 0.350-0.450(F)** | **0.420 (0.390-0.450)** | **0.440 (0.420-0.470)** | **0.390 (0.360-0.410)** | **0.000** |
| Decrease | 7/57 (12.3) | 4/32 (12.5) | 3/25 (12.0) |  |
| **MCV, fL; RI: 82.0-100.0** | **89.4 (86.6-92.4)** | **90.1 (87.8-93.4)** | **87.5 (85.5-90.0)** | **0.026** |
| Decrease | 2/57 (3.5) | 0/32 (0.0) | 2/25 (8.0) |  |
| **MCH, pg; RI: 27.0-34.0** | **31.0 (30.0-32.0)** | **31.7 (30.5-32.4)** | **30.3 (29.0-31.1)** | **0.001** |
| Decrease | 2/57 (3.5) | 0/32 (0.0) | 2/25 (8.0) |  |
| **MCHC, g/L; RI: 316-354** | **345 (342-350)** | **349 (344-352)** | **342 (338-345)** | **0.000** |
| Increase | 4/57 (7.0) | 3/32 (9.4) | 1/25 (4.0) |  |
| **RDW, %; RI: 11.00-16.00** | **11.50 (11.25-11.90)** | **11.50 (11.23-12.05)** | **11.60 (11.25-11.90)** | **0.809** |
| **PLT, $\times 10^9/L$; RI: 125-350** | **183 (161-229)** | **179 (156-219)** | **206 (166-256)** | **0.225** |
| Decrease | 7/57 (12.3) | 4/32 (12.5) | 3/25 (12.0) |  |

**Table 2 Laboratory findings of patients with COVID-19 at onset to hospital admission (continued)**
| **Laboratory parameters** | **All patients** | **Male** | **Female** | **P value** |
| --- | --- | --- | --- | --- |
| **MPV, fL; RI: 6.5-12.0** | **9.4 (8.6-10.1)** | **9.2 (8.3-9.9)** | **9.6 (8.7-10.1)** | **0.187** |
| **PDW, %; RI: 9.0-17.0** | **12.4 (10.5-14.3)** | **12.5 (10.5-14.3)** | **12.0 (10.6-13.8)** | **0.859** |
| Increase | 4/57 (7.0) | 2/32 (6.3) | 2/25 (8.0) |  |
| **PCT, %; RI: 0.108-0.282** | **0.180 (0.140-0.200)** | **0.160 (0.140-0.190)** | **0.200 (0.150-0.230)** | **0.034** |
| Increase | 4/57 (7.0) | 1/32 (3.10) | 3/25 (12.0) |  |
| Decrease | 3/57 (5.3) | 2/32 (6.30) | 1/25 (4.0) |  |
| **TT, s; RI: 14.0-20.0** | **15.6 (15.0-16.2)** | **15.2 (14.7-16.0)** | **15.9 (15.3-16.4)** | **0.029** |
| **APTT, s; RI: 22.2-38.0** | **34.7 (31.8-36.3)** | **35.5 (31.5-37.0)** | **33.6 (31.8-35.1)** | **0.075** |
| Increase | 4/37 (10.8) | 4/21 (19.0) | 0/16 (0.0) |  |
| **PT, s; RI: 11.0-13.0** | **11.9 (11.6-13.2)** | **12.8 (11.7-13.8)** | **11.7 (11.4-12.5)** | **0.043** |
| Increase | 9/37 (24.3) | 9/21 (42.9) | 0/16 (0.0) |  |
| Decrease | 3/37 (8.1) | 3/21 (14.3) | 0/16 (0.0) |  |
| **INR; RI: 0.90-1.30** | **1.07 (1.00-1.18)** | **1.10 (1.00-1.20)** | **1.01 (1.00-1.10)** | **0.080** |
| Increase | 3/37 (8.1) | 3/21 (14.3) | 0/16 (0.0) |  |
| **FBG, g/L; RI: 2.00-4.00** | **2.20 (2.01-3.24)** | **2.10 (2.02-3.07)** | **2.25 (1.93-3.43)** | **0.634** |
| Increase | 5/37 (13.5) | 2/21 (9.5) | 3/16 (18.8) |  |
| Decrease | 6/37 (16.2) | 2/21 (9.5) | 4/16 (25.0) |  |
| **hsCRP, mg/L; RI: ≤6.0** | **7.50 (2.4-32.1)** | **11.30 (2.8-33.6)** | **6.0 (2.0-20.5)** | **0.398** |
| Increase | 23/45 (51.1) | 15/27 (55.6) | 8/18 (44.4) |  |
| **Mb, ng/ml; RI: 0.0-61.5** | **22.7 (19.2-26.0)** | **21.9 (19.1-26.3)** | **22.9 (19.2-26.0)** | **0.901** |
| **cTnI, ng/ml; RI: <1.50** | **1.35 (1.10-1.88)** | **1.40 (1.10-1.80)** | **1.30 (0.95-2.16)** | **0.926** |
| Increase | 6/28 (21.4) | 6/15 (40.0) | 0/13 (0.0) |  |
| **CK, U/L; RI: 50-310(M), 40-200(F)** | **76 (55-126)** | **105 (62-152)** | **66 (43-77)** | **0.003** |
| Increase | 3/49 (6.1) | 3/28 (10.7) | 0/21 (0.0) |  |
| **CK-MB, U/L; RI: 0.0-24.0** | **15.0 (11.0-23.5)** | **13 (11.0-22.5)** | **18.0 (11.5-24.5)** | **0.412** |
| Increase | 11/49 (22.5) | 6/28 (21.4) | 5/21 (23.8) |  |
| **LDH, L; RI: 120-250** | **209 (184-265)** | **206 (186-279)** | **227 (164-261)** | **0.777** |
| Increase | 15/49 (30.6) | 8/28 (28.6) | 7/21 (33.3) |  |

**Table 2 Laboratory findings of patients with COVID-19 at onset to hospital admission (continued)**
| **Laboratory parameters** | **All patients** | **Male** | **Female** | **P value** |
| --- | --- | --- | --- | --- |
| **AST, U/L; RI: 15.0-40.0(M), 13.0-40.0(F)** | **25.00 (20.0-30.5)** | **24.0 (20.0-31.8)** | **26.0 (20.5-30.0)** | **0.840** |
| Increase | 8/49 (16.3) | 6/28 (21.4) | 2/21 (9.5) |  |
| **ALT, U/L; RI: 9.0-50.0(M), 7.0-40.0(F)** | **25.0 (19.0-46.0)** | **26.5 (16.3-57.8)** | **25.0 (19.5-42.5)** | **0.613** |
| Increase | 13/49 (26.5) | 8/28 (28.6) | 5/21 (23.8) |  |
| **GGT, U/L; RI: 10.0-60.0(M), 7.0-45.0(F)** | **24.0 (15.0-40.0)** | **33.5 (17.5-59.8)** | **19.0 (12.5-33.3)** | **0.024** |
| Increase | 9/48 (18.8) | 7/28 (25.0) | 2/20 (10.0) |  |
| **CHE, U/L; RI: 5000-12000** | **8171 (6671-9023)** | **8426 (6704-9110)** | **8034 (6610-8812)** | **0.563** |
| Decrease | 1/41 (2.4) | 1/23 (4.3) | 0/18 (0.0) |  |
| **TP, g/L; RI: 65.0-85.0** | **67.8 (65.0-71.7)** | **67.8 (65.2-71.9)** | **67.4 (65.0-72.0)** | **0.848** |
| Decrease | 11/49 (22.4) | 6/28 (21.4) | 5/21 (23.8) |  |
| **ALB, g/L; RI: 40.0-55.0** | **43.7 (42.1-45.7)** | **44.6 (41.7-47.0)** | **43.6 (42.2-44.7)** | **0.379** |
| Decrease | 5/49 (10.2) | 3/28 (10.7) | 2/21 (9.5) |  |
| **GLB, g/L; RI: 20.0-40.0** | **23.7 (21.5-26.8)** | **23.3 (20.5-26.3)** | **24.1 (22.1-28.9)** | **0.104** |
| Decrease | 6/49 (12.2) | 5/28 (17.9) | 1/21 (4.8) |  |
| **TBIL, µmol/L; RI: ≤23.0** | **7.9 (6.8-12.8)** | **8.8 (7.6-13.2)** | **7.1 (6.1-9.5)** | **0.009** |
| Increase | 1/49 (2.0) | 1/28 (3.6) | 0/21 (0.0) |  |
| **DBIL, µmol/L; RI: ≤8.0** | **3.6 (3.2 -5.1)** | **4.3 (3.6-5.2)** | **3.2 (2.7-3.9)** | **0.001** |
| Increase | 1/49 (2.0) | 1/28 (3.6) | 0/21 (0.0) |  |
| **IBIL, µmol/L; RI: 5.10-21.40** | **4.40 (3.30-7.00)** | **4.75 (3.63-7.88)** | **4.00 (2.95-5.20)** | **0.106** |
| Decrease | 29/49 (59.2) | 15/28 (53.6) | 14/21 (66.7) |  |
| **BUN, mmol/L; RI: 3.10-8.00 (M), 2.60-7.50 (F)** | **3.69 (3.03-4.70)** | **3.88 (3.10-4.77)** | **3.62 (2.91-4.34)** | **0.284** |
| Increase | 1/49 (2.0) | 1/28 (3.6) | 0/21 (0.0) |  |
| **Cr, µmol/L; RI: 57.0-97.0(M), 41.0-73.0(F)** | **68.0 (58.8-77.0)** | **74.4 (68.9-85.2)** | **57.5 (49.8-64.2)** | **0.000** |
| Increase | 2/49 (4.1) | 1/28 (3.6) | 1/21 (4.8) |  |
| **CO<sub>2</sub>, mmol/L; RI: 22.0-29.0** | **20.7 (18.9-22.5)** | **20.2 (18.8-23.1)** | **21.3 (18.8-22.4)** | **0.976** |
| Increase | 1/49 (2.0) | 1/28 (3.6) | 0/21 (0.0) |  |
| Decrease | 34/49 (69.4) | 19/28 (67.9) | 15/21 (71.4) |  |

**Table 2 Laboratory findings of patients with COVID-19 at onset to hospital admission (continued)**
| Laboratory parameters | All patients | Male | Female | *P* value |
| --- | --- | --- | --- | --- |
| **Glu, mol/L; RI: 3.89-6.11** | **6.17 (5.54-7.05)** | **6.44 (5.58-7.53)** | **5.75 (5.46-6.85)** | **0.312** |
| Increase | 25/49 (51.02) | 17/28 (60.7) | 8/21 (38.1) |  |
| **K, mmol/L; RI: 3.50-5.30** | **4.05 (3.78-4.37)** | **4.08 (3.82-4.40)** | **4.03 (3.72-4.37)** | **0.799** |
| Increase | 1/50 (2.0) | 0/28 (0.0) | 1/22 (4.5) |  |
| Decrease | 3/50 (6.0) | 2/28 (7.1) | 1/22 (4.5) |  |
| **Na, mmol/L; RI:137-147** | **138 (137-140)** | **138 (136-140)** | **139 (137-140)** | **0.148** |
| Decrease | 14/50 (28.0) | 11/28 (39.3) | 3/22 (13.6) |  |
| **Cl, mmol/L; RI: 99-110** | **100 (97-102)** | **98 (95-101)** | **101 (99-103)** | **0.037** |
| Decrease | 15/50 (30.0) | 15/28 (53.6) | 0/22 (0.0) |  |

Data with bold are median (*P*<sub>25</sub>-*P*<sub>75</sub>), and others are *n* (%); RI: reference intervals; M: male; F: female; *P* values: comparing male group and female group were from *Mann–Whitney U* test.

### Temporal trajectories of laboratory tests

The average value of EO was near the lower limit of RI at onset, while it continued to significantly increase but always below the upper limit of RI until discharge. The average value of NE was gradually increased from onset, peaking on day 12, and then steadily falling to the lower limit of RI. The average value of LY was approached the lower limit of RI for the first 5 days and then rose slowly thereafter, reaching peak around day 20. The ratio of NE/LY was peaked on day 5 and slowly declined thereafter. The average value of MO and BA showed u-shaped trend, peaking on day 17 and day 20, respectively. Platelet parameters—PLT, PCT and mean platelet volume (MPV) were within the RI in all cases. The average value of PCT were peaked on day 7 and slowly declined thereafter, while the average value of MPV was decreased sharply after day 7, even close to the lower limit of RI. There was no significant change in erythrocyte parameters, including RBC, HGB, HCT, MCH, and MCHC. However, the average value of MCV showed an upward trend from day 8. The average value of cTnI was sharply declined, and the average value of carbondioxide combining power (CO_2_) was significant increased throughout the course of the disease. The other haematological, biochemical, and immunological tests were not found significant trend over time. Interestingly, the average value of serum Na showed upward trend after day 15. The trajectories of laboratory tests over time were shown in Figure 2.

**Figure 2.**
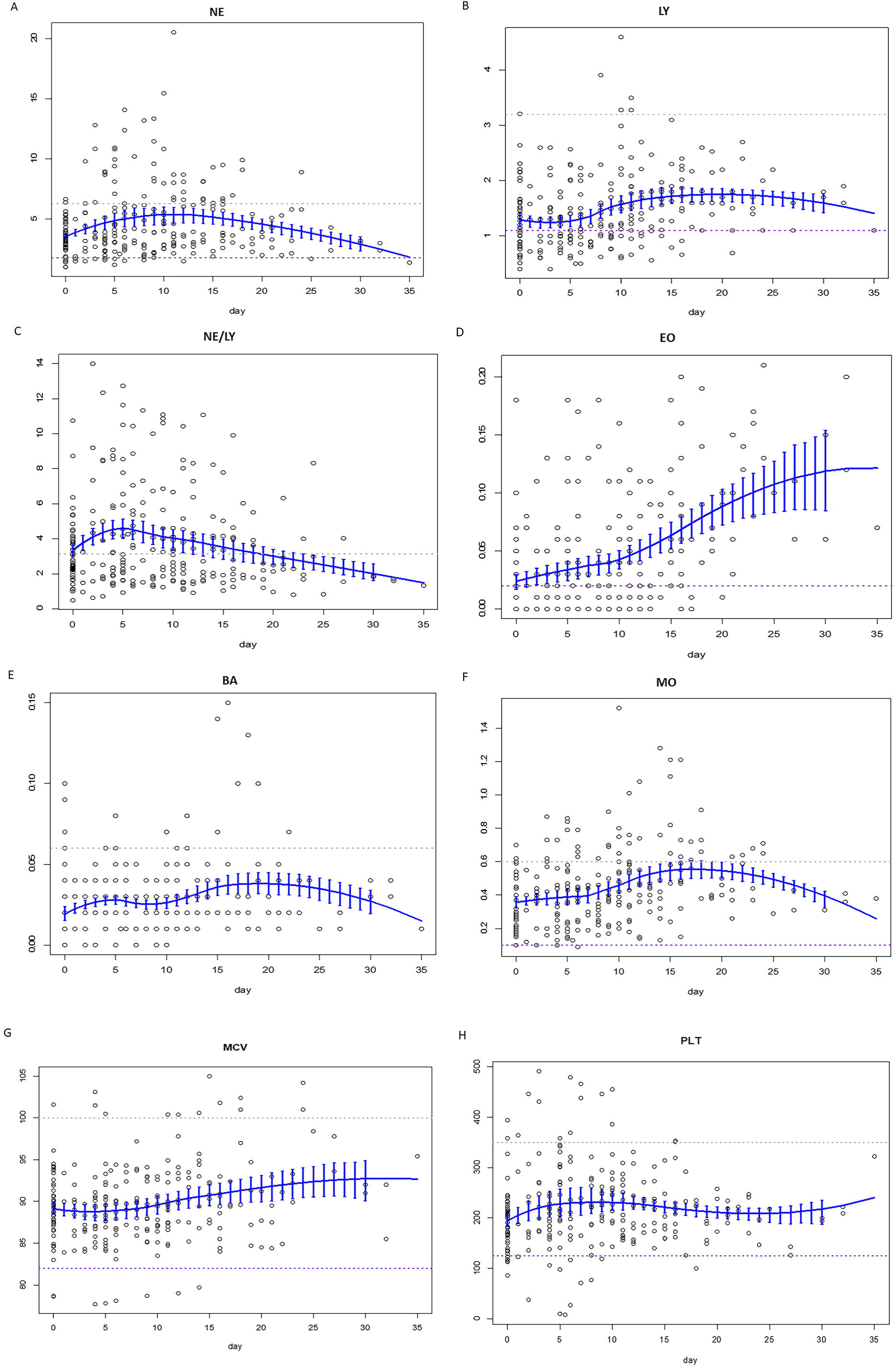
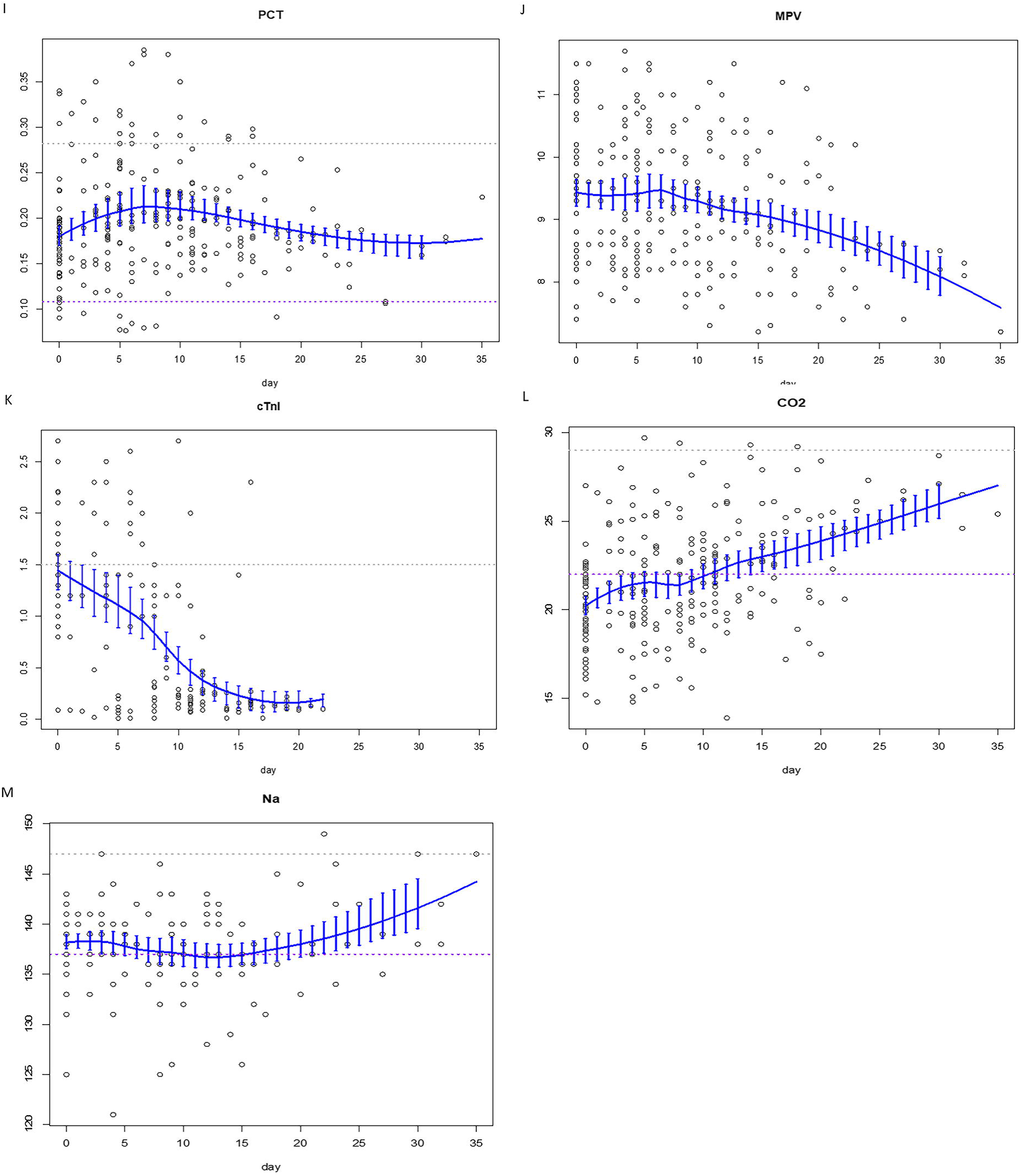
Temporal trajectories of laboratory tests. (A)NE. (B)LY. (C)NE/LY. (D)EO. (E)BA. (F)MO. (G)MCV. (H)PLT. (I)PCT. (J)MPV. (K) cTnI. (L)CO_2_. (M)Na. Here, only the representative parameters over time were presented. The gay dotted line represents the upper limit of RI whereas the purple dotted line represents the lower limit.

### Predictors of recovery

The measures of tests during the early period of hospitalized days (<5 days) were summarized their respective means, which were used in the hereafter analysis. Univariate Cox regression models indicated that PCT, PLT, EO, EO (%), MO (%) and CO_2_ met the inclusion criteria (i.e., *P*<0.1). The correlation plot of these 6 tests and age was presented in Figure 3A. As expected, PCT and PLT, and EO and EO (%) were highly positively correlated to each other. Then the refined feature selection step was carried out with these 6 tests, age and gender as candidate markers. While LASSO selected EO (%), age, PLT and CO_2_, backward selection chose CO_2_ and stepwise selection chose EO, PLT and CO_2_. The AUC under ROC curves at days 10, 14 and 21 for these three models were presented in Figure 3B as well. Basically, both model 1 and model 3 outperformed model 2. Since EO (%) and EO were highly correlated, age, EO, PLT and CO_2_ were included in the final model, and all markers but age was statistically significant (*P*<0.05). As shown by the nomogram of this final model (Figure 4), when PLT and CO_2_ at the early days were low the chance of quick recovery decreased, whereas when EO at the early days were low the probability of quick recovery increased.

**Figure 3.**
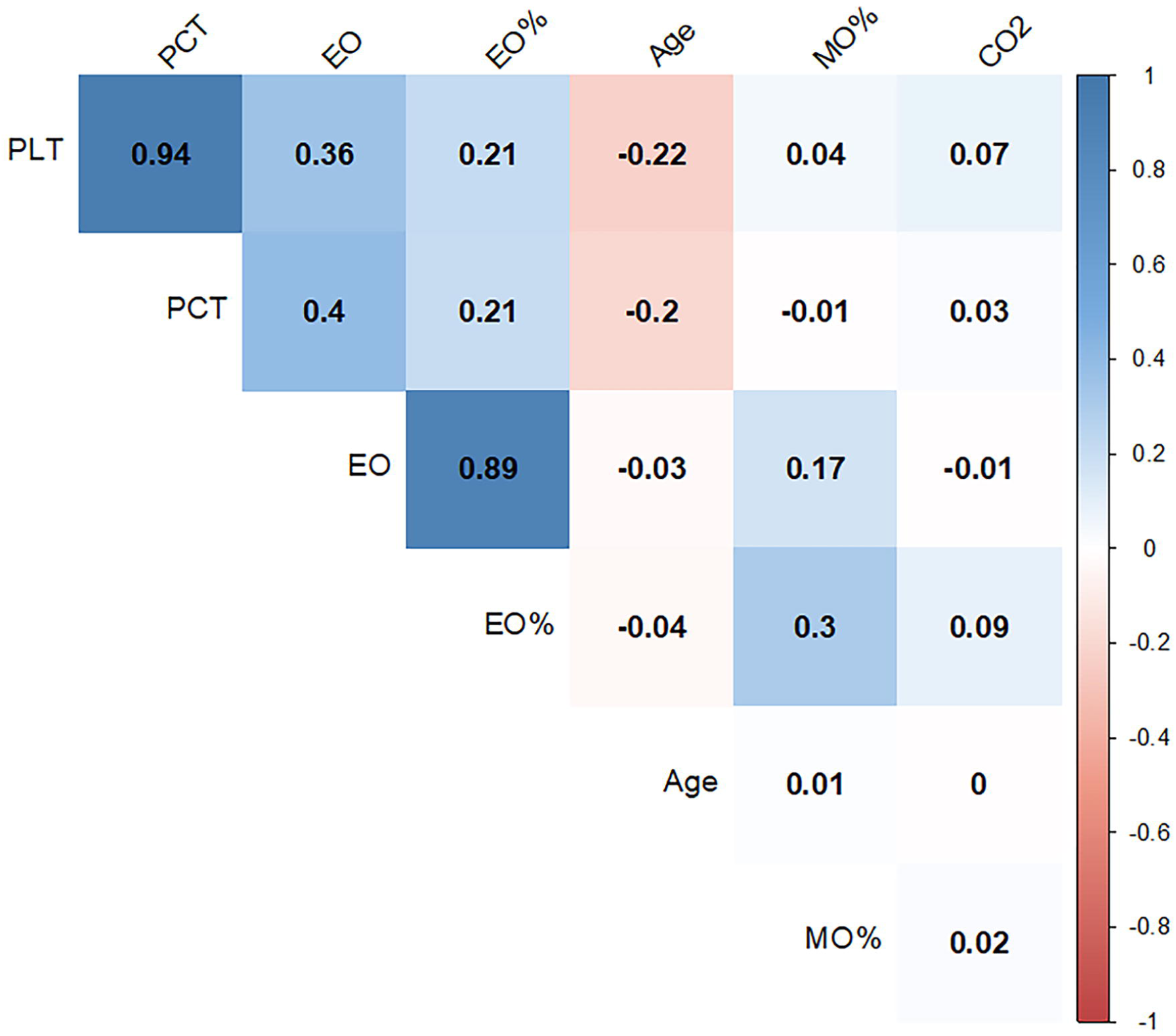
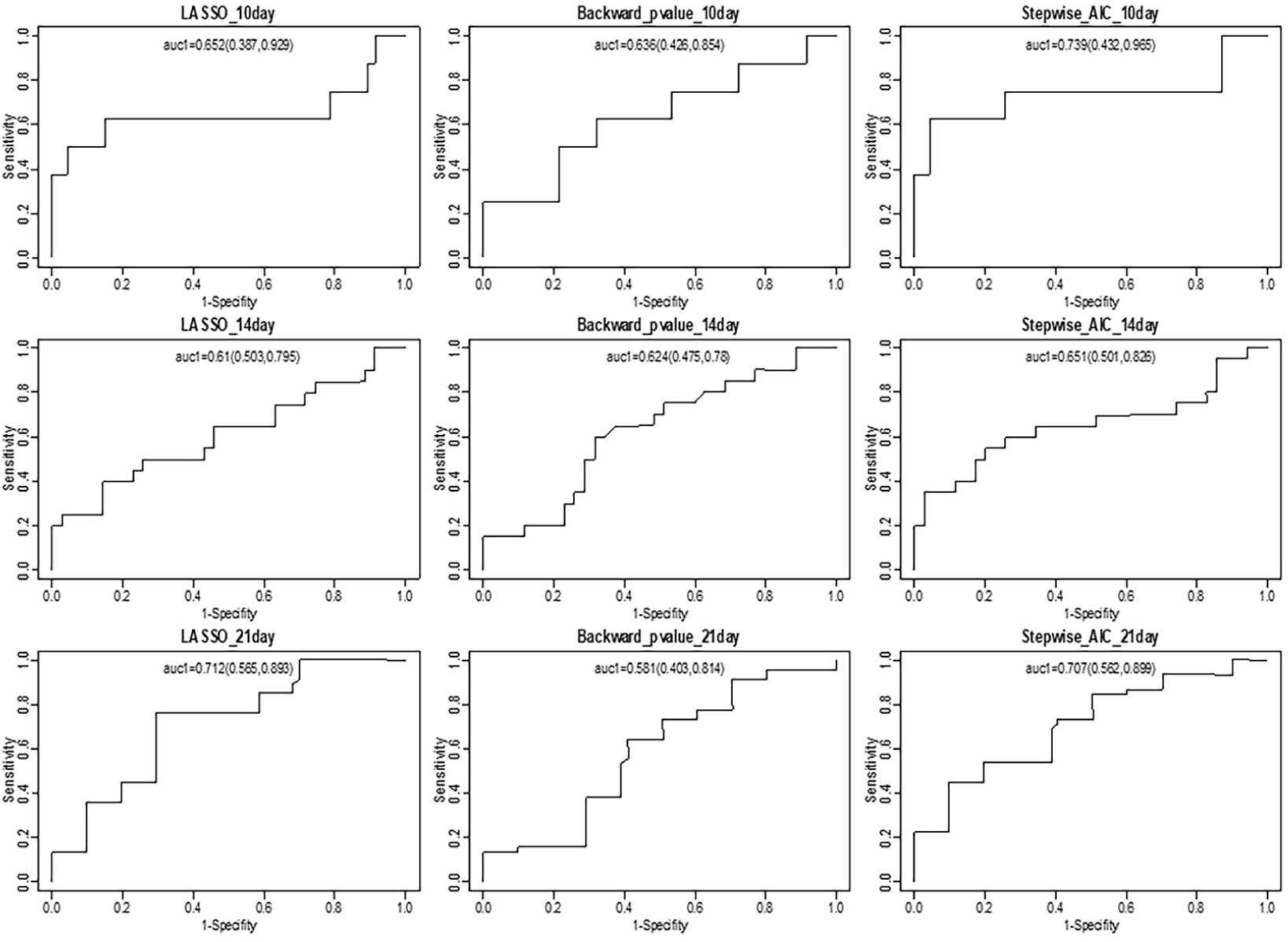
Feature selection process of relevant laboratory tests that have prognostic values for recovery. (A) correlation plot of selected tests by univariate Cox regression models and age. (B) Area under curves (AUC) of ROC for three feature selection models at days 10, 14 and 21. Here, LASSO, backward selection and stepwise selection were used.

**Figure 4.**
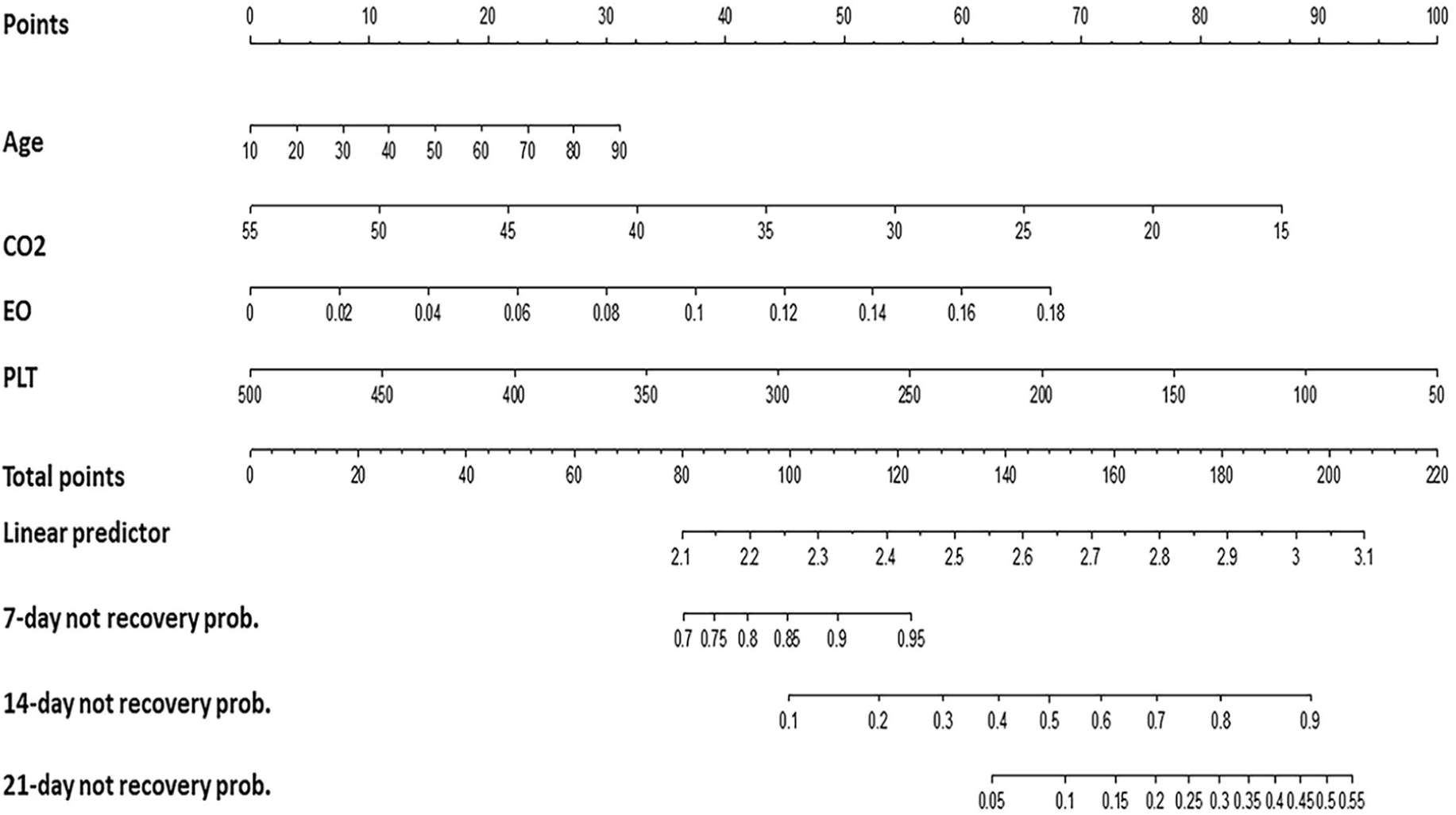
Nomogram to illustrate how age, EO, PLT and CO_2_ at early days of admission to hospitals (<5 day) are related to recovery.

## Discussion

Of all 59 cases diagnosed as COVID-19 in the three designated tertiary hospitals in Jilin province, most of them were middle and elderly aged. In the present study, at the final date of follow-up, all the patients were recovered and discharged. Of the 59 cases, imported, primary and secondary cases accounted for 44.1%, 42.4% and 13.6%, respectively. In the present study, the 3 patients with severe type were all over 45 years old.

### Haematological tests

In the present study, eosinopenia was shown in 52.6% cases at onset, similar with the recent research.(7) As shown in its trajectory (Figure 2), the level of EO continued to increase thereafter until discharge. Therefore, the longitudinal change patterns of EO could be used as an early predictor for COVID-19. For the cases with typical clinical manifestations and changes of chest CT with and without lymphopenia, eosinopenia was predictive for the diagnosis. For the confirmed cases, continuous rise of EO value may be a sign for recovery.

The value of LY in 40.4% cases was declined at onset in the present study, consistent with other reports.^(7, 8)^ Lymphopenia showed that SARS-CoV-2 may mainly act on lymphocytes, as does coronavirus. The average value of LY was slowly elevated after day 5 with peaking around day 20, as well as peaking on day 5 and declining thereafter for the ratio of NE/LY, indicating the gradual return of number and function for lymphocytes in recovered patients. In a recent study, the incidence of severe illness was 9.1% in age ≥50 and NE/LY < 3.13 patients, and half of patients with age ≥ 50 and NE/LY ≥ 3.13 would develop severe illness.(9) In the present study, the average ratio of NE/LY was higher than 3.13 at onset with development of severe type in 3 cases, indicating the certain value of the ratio of NE/LY.

The value of PLT was under the lower limit of RI in 12.3% cases at onset, similar with the recent research of 12% cases (*n*=99) in a hospital,(8) but significantly lower than the report of 36.2% cases (*n*=1099) in 552 hospitals in China.(10) Thrombocytopenia was more significantly in the patients with severe type and critical type. In the present study, the average value of PCT was peaked on day 7 and slowly declined thereafter, while the average value of MPV was decreased sharply after day 7. These longitudinal change patterns indicated platelet parameters may be predictors for recovered patients.

The erythrocyte parameters-counts of RBC, HGB, HCT, MCV, and MCH were all less than 15% below the lower limit of RI at onset. However, the average value of MCV showed an upward trend from day 8, indicating constant recovery thereafter.

Based on these temporal trajectories of key laboratory tests (Figure 2), we speculate that with medical intervention, patients would experience an improvement approximately after one week at hospital stays. Further investigation is warranted to test if day 7 or 8 is a turning point for recovery with medical intervention.

### Biochemical and immunological tests

Acute cardiac injury was found in approximately 12% patients with COVID-19.(1) In the present study, the values of AST, LDH, CK, CK-MB, and cTnI, serum cardiac markers, were beyond the upper limit of RI from 6.1% to 30.6% at onset. Consistent with the report,(11) cardiac injury is a common condition among hospitalized patients with COVID-19, and it is associated with higher risk of in-hospital mortality. Therefore, it is necessary to actively prevent and respond to myocardial injury, reduce the possibility of irreversible remodeling of the myocardium.

Hepatic injury was also found in the present study. The values of ALT, GGT, TBIL, and DBIL were beyond the upper limit of RI from 2.0% to 26.5%, while the values of CHE, TP, ALB, GLB, and IBIL were below the lower limit of RI from 2.4% to 59.2%. Liver function tests indicate gently hepatic injury in the patients with COVID-19. Although kidney injury was not severe, the values of BUN and Cr above the upper limit of RI were also found in 2.0%, and 4.1% cases, respectively. The disturbance of electrolyte balance was also a common condition in the patients with COVID-19, with decreased value of K, Na and Cl from 6.0% to 30.0%.

### Prognostic factors of recovery

In the study, age, EO, PLT and CO_2_ were selected as the useful prognostic factors affecting the prognosis of recovery by LASSO, backward and stepwise. The nomogram was established base on age, EO, PLT and CO_2_, which were used to predict the recovery rates of 7-day, 14-day, and 21-day. We suggest that patients with COVID-19 can improve risk stratification and management according to age, EO, PLT and CO_2_ model.

### Limitations of this study

Disadvantages are as follows: (1) there were 59 cases enrolled in the present study and the sample size may be not very large. Nevertheless, given that Jilin province is the least affected area in China, this study covered approximately 70% of diagnosed cases in Jilin province, in addition to longitudinal measurements of laboratory tests had been collected, these rendered the sample size of this study adequate for reliable statistical estimations; (2) the data of clinical manifestation in the patients with COVID-19 was not included since this study mainly focused on the temporal trajectories of laboratory findings.

## Conclusion

The longitudinal change patterns of key laboratory tests in the patients with COVID-19 were found. The average value of EO continued to significantly increase, while the average value of MPV was decreased sharply after day 7. Age, EO, PLT and CO_2_ may used to predict the recovery probability. Risk stratification and management could be improved for the patients with COVID-19 according to temporal trajectories of laboratory tests.

## Data Availability

The raw data required to reproduce these findings cannot be shared at this time as the data also forms part of an ongoing study.

## Acknowledgements

None.

## Ethical approval

This study was approved by the Ethics Committee of the First Hospital of Jilin University, Changchun Infectious Disease Hospital and Siping Infectious Disease Hospital.

